# Measuring Italian Citizens’ Engagement in the First Wave of the COVID-19 Pandemic Containment Measures: A Cross-sectional Study

**DOI:** 10.1101/2020.04.22.20075234

**Authors:** Guendalina Graffigna, Serena Barello, Mariarosaria Savarese, Lorenzo Palamenghi, Greta Castellini, Andrea Bonanomi, Edoardo Lozza

## Abstract

**Background:** In January 2020, the coronavirus disease 2019 (COVID-19) started to spread in Italy. The Italian government adopted urgent measures to hold its spread. Enforcing compliance to such measures is crucial in order to enhance their effectiveness. Engaging citizens in the COVID-19 preventive process is today urgent in Italy and around the world. However, to the best of our knowledge, no previous studies have investigated the role of health engagement in predicting citizens’ compliance to health emergency containment measures.

**Method:** An online survey was administered between February 28th and March 4th 2020 on a representative sample of 1000 Italians. The questionnaire included a measure of Health Engagement (PHE-S) and a series of ad hoc items intended to measure both affective and behavioral responses of the citizens to the emergency in terms of perceived susceptibility to and severity of the disease, orientation towards health management, change in habits and in purchases. To investigate the relationship between Health Engagement and these variables, a series of ANOVAs, Logistic regressions and crosstabs have been carried out.

**Results:** Less engaged people show higher levels of perceived susceptibility to the virus and of severity of the disease; they trust less scientific and healthcare authorities, they feel less self-effective in managing their own health - both in normal conditions and under stress - and are less prone to cooperate with healthcare professionals. Low levels of Health Engagement are also associated with a change in the usual purchase behavior.

**Conclusions:** The Patient Health Engagement Model (PHE) provides a useful framework for understanding how people will respond to health threats such as pandemics. Therefore, intervention studies should focus on particular groups and on raising their levels of engagement to increase the effectiveness of educational initiatives devoted to promote preventive behaviors.

## 1. Background

In January 2020, the coronavirus disease 2019 (COVID-19) caused by severe acute respiratory syndrome coronavirus 2 (SARS-CoV-2) started to spread in Italy. As of March 17th, 2020, a total of 31.506 COVID-19 cases with 2.503 deaths and 2.941 recovered had been reported in Italy. On Jan 30, 2020, the World Health Organization (WHO) declared the coronavirus outbreak a public health emergency of international concern (PHEIC). Since March 8th, 2020, the Italian government adopted very urgent and restrictive measures to hold the virus spread and its potential impact on the population. Several cities - identified as “red areas” - have been put under quarantine, hoping to stop the disease from spreading to other parts of the country. This situation is globally unprecedented at least for two main reasons. First, to control of the COVID-19 outbreak, the governmental authorities have suddenly adopted very extreme public measure such as locking down cities, deeply reorganizing healthcare services to cope with the rapid increasing demand for acute care, imposing school and university closure, suggesting - where possible - smart-working solutions and transportation restrictions, deploying thousands of healthcare workers to more heavily affected regions, and running wide public health messaging campaigns for consumers’ education. Second, consumers are overwhelmed by rather mixed and confounding information, partly because scientific discovery related to COVID-19 is constantly evolving with the course of the disease outbreak, partly due to the rapid increase in misleading or false news. Therefore, all these measures are currently having a deep impact on Italian people’s attitudes, daily habits and consumer behaviors.

Like in other similar situations, prior to the availability of an effective vaccination therapy, strategies to mitigate and control the impact of the pandemic typically involve “non pharmacological” interventions (1,2), and lever on citizens autonomous responses to public health preventive measures. In particular, past literature suggested as people appear to respond to an epidemic by voluntarily undertaking specific behaviors in order to protect themselves (3,4). However, in some cases, these behaviors may not correspond to an objective evaluations of risk (5,6), but depend also on individual subjective evaluations, thus becoming potentially counterproductive. For this reason, there has been a rapid rise in interest in understanding the determinants of people’s behavioral change that may influence the adequacy of the response to health emergencies (7,8). People dealing with these situations, indeed, may experience negative attitude, feelings of uncertainties and alarmism (9). These reactions might potentially end in risky habits and inadequate and disorganized behaviors, both for individuals and community (10,11), affecting public health outcomes. Therefore, to study the subjective factors implied in such reactions is of much relevance for effectively sensitize the general public and identify high-risk targets (12). Along with structural and immutable factors such as socio-demographics, scholars have previously attempted to understand the subjective determinants of citizens’ changing attitudes and behaviors in a pandemic emergency. In particular, authors identified the risk perception as one of the most relevant variable in determining citizens’ response to global pandemic disease (13,14), in relation to a series of factors such as the perception of economic impact (15); efficacy believes related to health (16); level of literacy and knowledge elaboration (17,18). Another important factor identified is the level of subjective anxiety, which influences both citizens’ attitudes towards the emergency disease and consequent preventive behaviors (19–21). Other subjective factors accounted for the change peoples’ habits in pandemic emergencies are also the ones related to the perceived effect of one individual behavior, such as perceived costs and benefits of preventive behaviors on the disease spread (22,23) or perceived impact of an individual’s behavior on other individual’s outcomes (21,24).

Among other variables accounting for a change in citizens’ attitudes, habits and behaviors, recently scholars have showed the role of health engagement in affecting health-related behaviors and preventive habits (25–28). More in details, people with high levels of health engagement have been identified as more effective in adopting behavioral change suggestions and in adhere to medical prescriptions (29–31). However, previous literature has also demonstrate how individuals may be in different phases of their process of engagement (26,28), thus resulting more or less ready to enact a change in their way to cope with a critical events and to comply to prescribed preventive conducts.

In the current COVID-19 outbreak in Italy – as well as in other countries – citizens are experimenting a life-threatening situation, leading to profound changes in attitudes, habits and behaviors, also potentially negative for consumers’ health and virus containment. Making citizens aware about their crucial role in avoiding the rapid spread of the virus and engaging them in the COVID-19 preventive process is today urgent in Italy and around the world.

However, to the best of our knowledge, no previous studies have investigated the role of people health engagement in determining citizen attitudes, habits and compliance to containing measures in the occasion of an health emergency; moreover, previous literature have highlighted the need to apply a validated theoretical framework to the study of these phenomena in order to effectively predict people responses to the event and adherence to prescriptions (8).

For these reasons, we propose a study aimed at understanding citizens’ attitudes and behavioral responses to the current spread of COVID-19 in Italy and how they changed their daily habits and behaviors according to their level of health engagement. Results of this study will contribute to informing public health communication and targeted consumer educations activities.

### 1.1 Theoretical background

The Patient Health Engagement model (26,32) is a psychological framework which theorizes how individual health engagement results from a continuous emotional and motivational reframing of individuals own role perception in the management of a disease (i.e. from passive user of services to active partner of the healthcare system). According to this model, to become engaged means to be emotionally resilient and able to adjust to the health risks and specific requirements. This model also features peculiar ways of coping with health crisis, as can be considered the Covid-19 today. In particular, the model features four positions: the first position (“blackout”) is of complete disengagement, typically occurring when people feel vulnerable and without control over the perceived risk, psychologically frozen and behaviorally paralyzed. It follows the psychological position of “Arousal,” in which people have acquired an initial awareness about their actual situation of health risk but don’t have still enough knowledge and skills to manage it. They do not accept the impact of preventive requirements on the modification of their daily habits and appear hyper-vigilant over their body signals, iperactive and confused when seeking information on the health situation. Each unexpected news or change in the epidemic situation causes emotional alert and overwhelming emotional responses, with disorganized actions and behaviors. When individuals succeed in the process of emotional regulation and coping with the stressful condition, they achieve a position of Adhesion. In this phase, patients have matured a good psychological adaptation to the critical situation and appear able to manage their psychological di-stress connected to health emergency. They appear more motivated to comply with medical and preventive prescriptions. In this phase, moreover, patients acquire further skills to effectively managing their risk condition. Finally, when people mature a complete awareness of the characteristics and consequences of the critical situation, and assume a better responsible position in their behaviors and risk management they reach the “eudaimonic project” phase, which features a better, positive and optimistic approach to the situation, with an increased ability to deal with the uncertainty of the moment and a strong motivation to psychologically achieve the sense of a “new normality” (Figure 1).

**Fig. 1.**
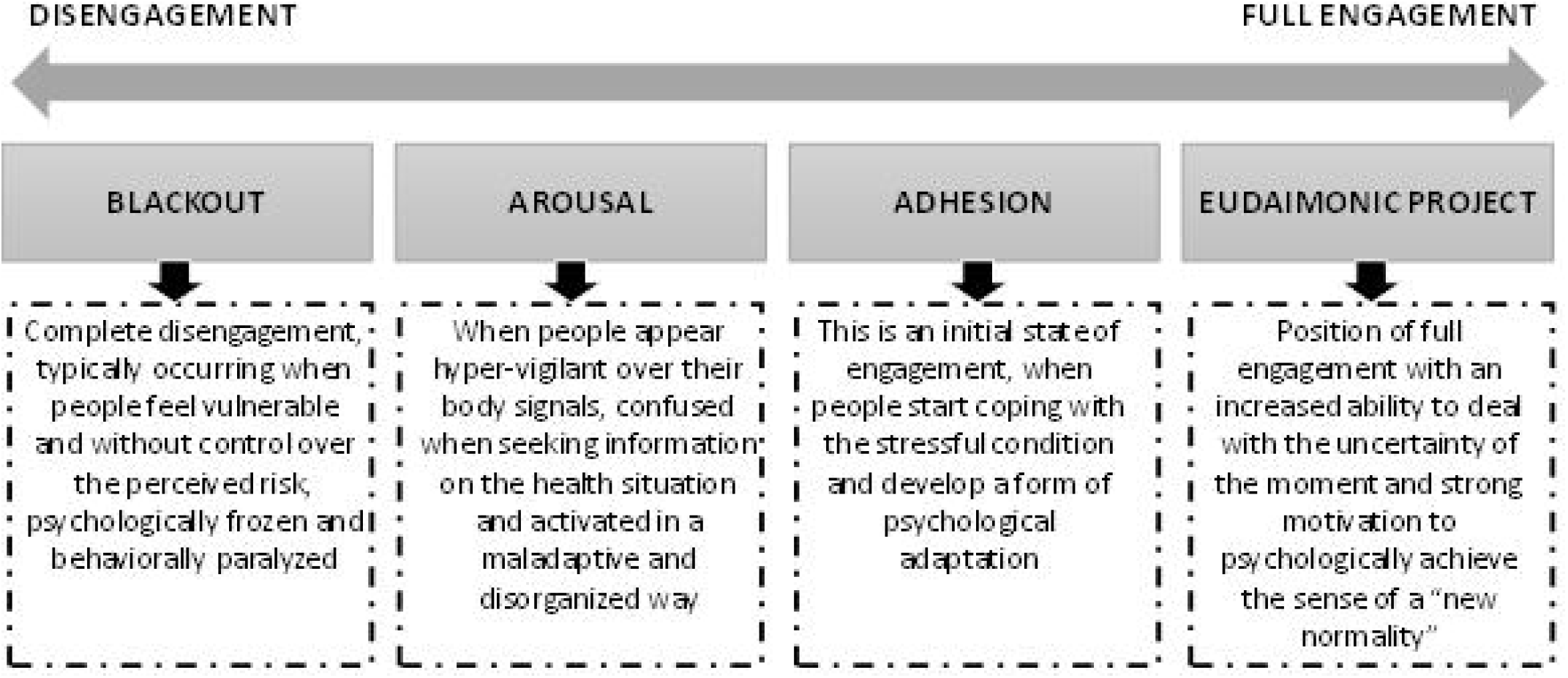
The Patient Health Engagement Model

### 1.2 Research questions

The study has the general aim to seek to explore how different levels of Health Engagement correspond to unique patterns of people reactions to the COVID-19 health emergency.

In particular, the study was guided by two main hypotheses:

1. Individuals with different levels of Health Engagement show different attitudinal responses towards the emergency. In particular:
  i. less engaged people perceive themselves more susceptible to the infection and afraid by the contagion than people with a higher Health Engagement profile;
  ii. different Health Engagement profiles show different attitudes to prevention and health management and different levels of trust towards authorities, scientific research and media during emergency periods;
2. Health Engagement is associated with changes in consumer behaviors and habits.

Results from this study are intended to provide evidence-based information that furthers our theoretical understanding of consumer reactions to pandemics outbreaks and might enables government to develop better and more effective policy responses to this and future pandemics.

## 2. Methods

### 2.1 Study design and participants

To investigate our hypotheses, a cross-sectional study was carried out between February 28th and March 4th 2020. A sample of 1000 Italians, representative of the Italian population for gender, age, employment, geographical area and dimension of urban center of residence, resident in all the different regions of Italy and over 18 years old was involved in the self-completion of a web questionnaire. Individuals were randomly selected from the consumers panel managed by Norstat srl (a company specialized in collecting data; (https://norstat.it/) using random digit dialing, to fill an online survey. Eligibility criteria for being involved in the study were purposefully kept minimal to make the results broadly applicable and only included being 18 years old or older, being able to read and understand Italian and live in Italy. People belonging to the online panel were carefully screened for authenticity and legitimacy via digital fingerprint and geo-IP-validation from the panel provider. In this study, in order to guarantee data quality, respondents were asked to confirm their demographics. From the 1000 recruited subjects, 32 were excluded because demographic data provided by the respondent and those provided from the panel were inconsistent (there were discrepancies between reported and known gender and/or age). Statistical analyses were hence carried out on a dataset composed of the answers of 968 respondents. All analyses have been carried out with IBM SPSS 23 (release 23.0.0.0).

Data collection is part of a broader longitudinal survey on consumers’ health engagement and behaviors. Ethical approval was obtained from the institutional review board at Catholic University of Milan (IRB#2019-12).

### 2.2 Study measures

After recruitment and consent, consumers were asked to complete an online survey. involving questions about health engagement, affective response and behavioral responses to the COVID-19 pandemic.

In particular, in order to answer the research questions, the survey was focused involved the measurement of the following variables:

- ***Health Engagement***: participants were asked to fill a revised version of the Patient Health Engagement Scale (PHE-s®). This measure, developed according to the Patient Health Engagement model (32), assesses the consumers health engagement level, defined as the “*people psychological readiness and sense of mastery to become active player in their own health management and health risk prevention*”. Previous studies demonstrated its robust psychometric proprieties (26). This scale features five ordinal items reflecting the continuum of engagement described in the four levels of the PHE model. According to the ordinal nature of the PHE-s®, the median score is considered the more reliable index to calculate the final patients’ scoring (26). According to the score obtained, each respondent result in one of the four levels of health engagement as described in the PHE model (i.e. blackout, arousal, adhesion, eudaimonic project). The scale is based on the assumption that the score obtained by the person should reflect his/her actual health engagement level. For this study purposes the PHE-s® was slightly revised in order to adapt the items formulation to the specific contest of the health emergency. For this reason, the psychometric characteristics of the revised version were tested.
- ***Attitudinal response towards to the COVID-19 health emergency***, involving:
  - In light of studies on risk processing (33), two elements of risk judgment were measured: *(a) risk severity: perceived potential severity of COVID-19 infection for their own health:* participants answered a question regarding how concerned they are about the emergency ranging from 1 (not concerned at all) to 10 (very concerned); *and (b) risk susceptibility: perceived likelihood to get COVID-19:* participants were asked to rate from 1 (very little) to 5 (a lot) their perceived risk of being infected by the new COVID-19 virus;
  - *orientation towards health management:* participants were asked a series of questions regarding their ability to manage their own health autonomously. In particular, they were asked how much they agreed to the following statements on a scale ranging between 1 (completely disagree) to 5 (completely agree):
    ▪ *self-responsibility*: “I am the responsible of diminishing my own risk of being infected by the new COVID-19 virus”
    ▪ *self-efficacy in health management*: “I can manage my own health effectively”
    ▪ *self-efficacy in stress management*: “I can manage my own health even when I’m distressed”
    ▪ *value of partnership in healthcare*: “it’s important to cooperate with healthcare professionals in defining how to manage my own health”
  - *trust in authorities:* participants were asked a series of questions regarding their trust in scientific research and the National Healthcare System. In particular, they were asked how much they agreed to the following statements on a scale ranging between 1 (completely disagree) to 5 (completely agree):
    ▪ *Trust in science*: “I fully trust scientific research”
    ▪ *Trust in NHS*: “I fully trust the National Healthcare System”
    ▪ *media reliability*: It is hypothesized that media tends to sensationalize food safety hazards. Participants were asked to respond to the following question: “I think that the emergency regarding Covid-19 has been created by an overblown mass-media hype”
- ***Behavioral responses*** involving:
  - *Information seeking:* participants were asked how much they were accessing information regarding COVID-19 on a series of both traditional and new medias (TV, newspapers, social networks, scientific journals etc.). Each media was rated from 1 (little) to 5 (a lot). Scores were then summed and an average was calculated, in order to obtain an indicator of how much a certain subject was searching for information regarding the virus;
  - *Consumer habits and purchasing behaviors:*
    - A series of four dichotomous, yes/no questions were asked regarding changes in consumer habits:
      ▪ *Reduced restaurant meals*: “have you reduced meals in restaurants?”
      ▪ *Reduced ethnic restaurant meals*: “have you reduced meals in ethnic restaurants?”
      ▪ *Products from the “red zones”*: “are you willing to buy food products coming from “red” zones?”
      ▪ *Stockpiling*: “have you -if you are the main responsible in food provisioning in your family-been stockpiling food and first need products?”
    - Changes in purchasing behaviors were also assessed with a series of questions surveying whether the buying of different products had diminished/remained the same/increased. In particular, the purchase of the following products was assessed:
      ▪ Fresh food (veggies, fruit, meat…)
      ▪ Frozen food
      ▪ Canned food
      ▪ Personal disinfection
      ▪ Personal care
      ▪ House disinfection
  - **Sociodemographic variables:** a series of socio-demographical data were also collected, including: age, gender, education and employment in order to characterize our sample. Moreover, participants were also asked to state whether they suffer from any chronical condition. Finally, participants were asked to state their region of residency. People were considered as “Coming from the red zones” if they stated to live in either Lombardia, Piemonte or Emilia-Romagna, Italian’s most involved regions at the time of data collection.

## 3. Results

### 3.1 Sample

Male participants were 473 (48.9%). Mean age was 44 years (SD = 14; range 18-70). For a more detailed description of the study sample see Table 1.

**Table 1.**
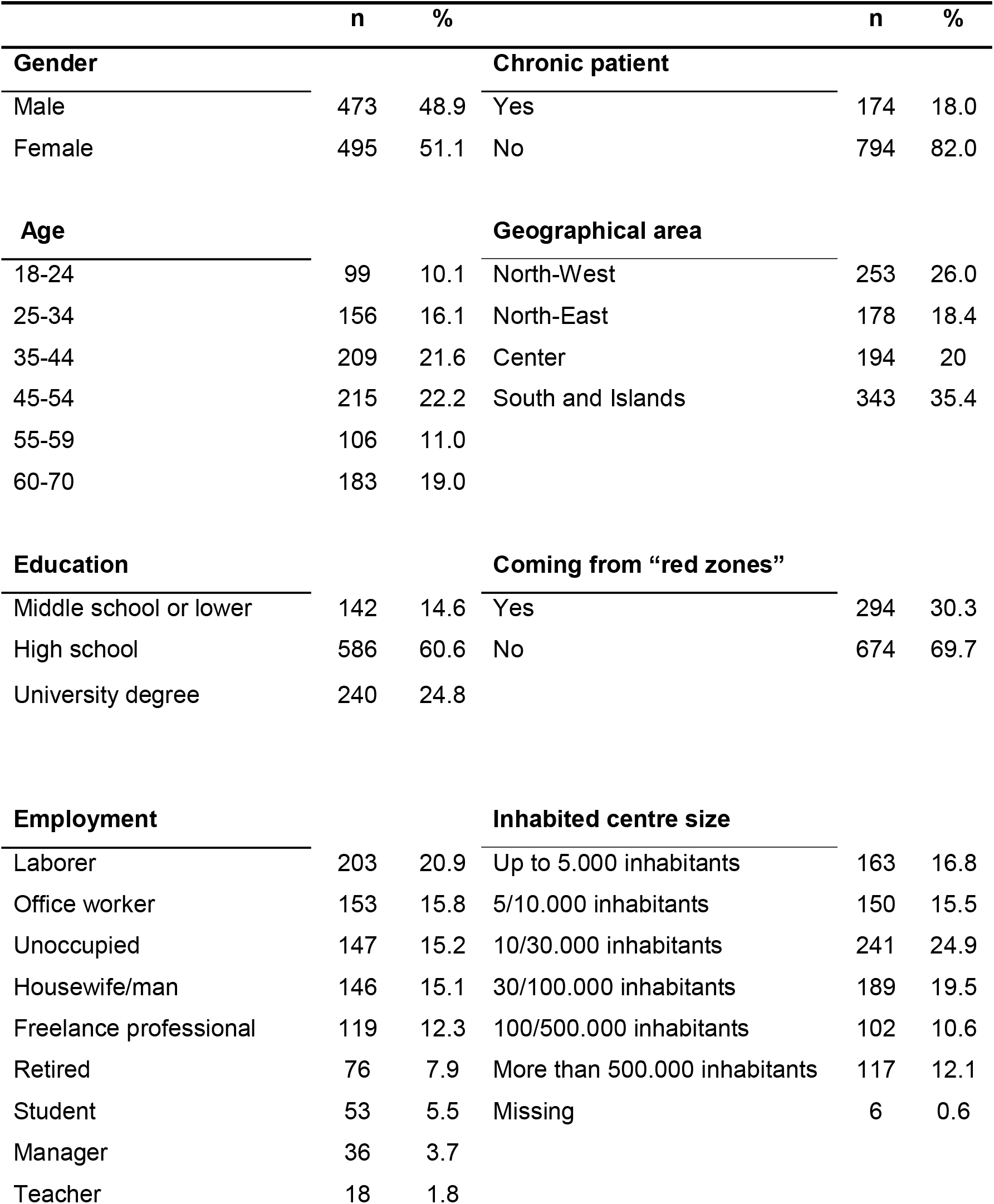
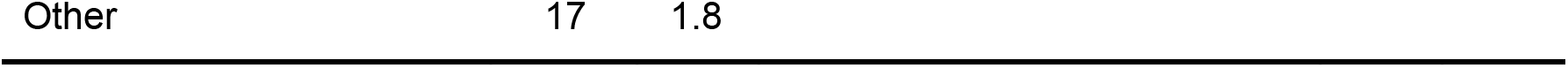
Demographic profiles of the sample (N = 968).

### 3.2 Psychometric proprieties of the PHE-s^®^ revised version

#### 3.2.1 Data Analysis

To evaluate the psychometric properties of the revised PHE-s^®^ scale, a Partial Credit Rasch Model (PCM) was performed to check uni-dimensionality and the fit of each item at the construct of interest. In particular, to check whether the items fitted the expected model, two items fit mean square (MNSQ) statistics (Infit and Outfit) were computed. If the data fit the Rasch model, the fit statistics should be between 0.6 and 1.4 (34). Analyses of difficulty and step parameters were conducted to guarantee a sufficient ranking of the different categories of response and to respect the monotonic order. The internal consistency of the items of the revised PHE-s^®^ was assessed using Ordinal Alpha via Empirical Copula Index (35) since the ordinal nature of the items. A reliability index superior to 0.7, 0.8, 0.9 can be interpreted as acceptable, good and excellent, respectively (36).

Finally, a Confirmatory Factor Analysis (CFA) was performed. Goodness-of-fit indexes (i.e. comparative fit index CFI, root mean square residual RMR and root mean square error of approximation RMSEA) were evaluated. A CFI > .90 was considered a good model fit (37), a RMR <.05 is desirable (38), whereas a RMSEA < .08 indicated an acceptable fit (39).

#### 3.2.2 Results

Table 2 shows the results of the Rasch Analysis.

**Table 2.**
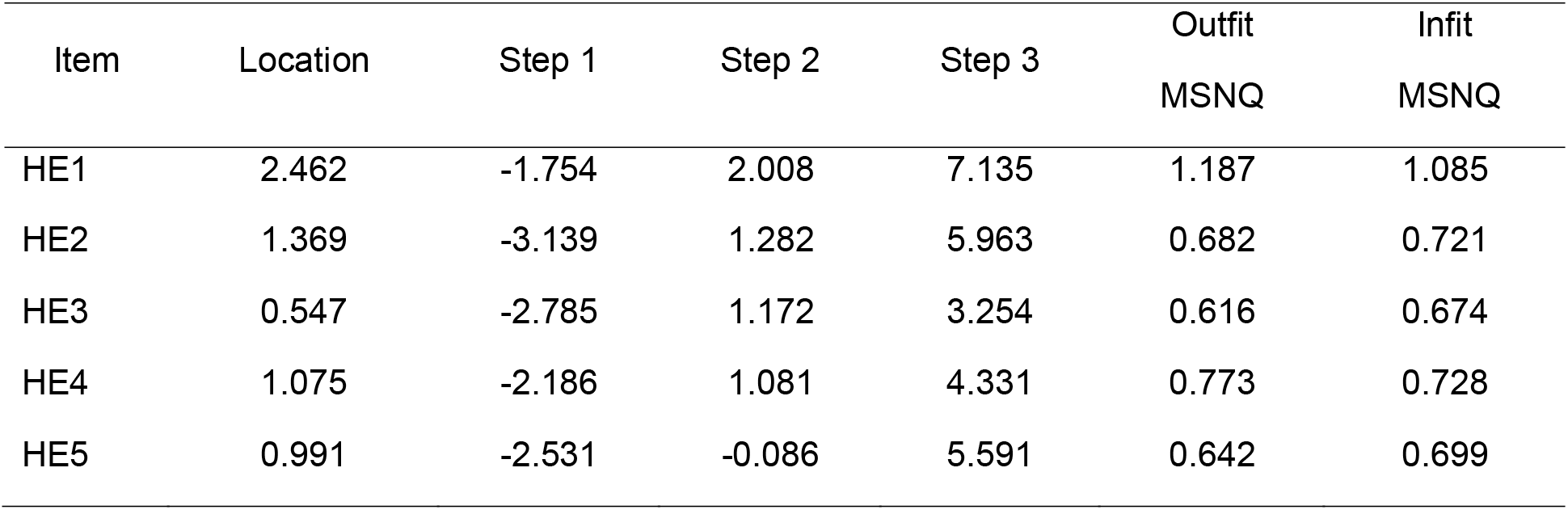
PCM and Item Fit Statistics

| Item | Location | Step 1 | Step 2 | Step 3 | Outfit<br>MSNQ | Infit<br>MSNQ |
| --- | --- | --- | --- | --- | --- | --- |
| HE1 | 2.462 | -1.754 | 2.008 | 7.135 | 1.187 | 1.085 |
| HE2 | 1.369 | -3.139 | 1.282 | 5.963 | 0.682 | 0.721 |
| HE3 | 0.547 | -2.785 | 1.172 | 3.254 | 0.616 | 0.674 |
| HE4 | 1.075 | -2.186 | 1.081 | 4.331 | 0.773 | 0.728 |
| HE5 | 0.991 | -2.531 | -0.086 | 5.591 | 0.642 | 0.699 |

The item statistics ranged from .674 to 1.085 for the infit MSQ and from .616 to 1.187 for the outfit MSQ. These values indicate an acceptable fit of the Rasch Model. The distances between subsequent thresholds showed acceptable distinction between the response options and measurement model fit. The Person Separation Index (PSI) was calculated to evaluate the reliability in the Rasch Model (PSI = .851). Rasch Model confirmed the unidimensionality of revised PHE-s^®^ scale and the fit of each item of the scale to the data.

The revised PHE-s^®^ had a quite good internal consistency, since the value of the Ordinal Alpha via Empirical Copula was equal to .788. Each item contributed significantly to the revised PHE-s^®^ scale score. So, the internal consistency of the revised PHE-s^®^ was satisfactory.

CFA showed reasonable goodness of fit indices. The fit indices met the criteria of fit for the hypothesized one-factor structure. All goodness of fit indices (CFI=0.994, RMR=0.008, RMSEA=0.066) suggested that the model is coherent with the data. The analysis of modification indices did not find any relation between the error covariance of the items. All the standardized to factor loadings are ranged from .532 to .820.

### 3.3 Descriptive statistics

There were no missing data in our dataset. For dichotomous and multiple-choice questions, answer frequencies and “I don’t know” answers are reported -where provided-in Table 3. However, in the following analyses “I don’t know” were considered as missing values. For other variables, descriptive statistics are reported in Table 4.

**Table 3.** Frequency distribution of items

|  | n | % |  | n | % |
| --- | --- | --- | --- | --- | --- |
| **Health Engagement Level** |  |  | **Products from the “red zones”** |  |  |
| Blackout | 11 | 1.1 | Yes | 498 | 51.1 |
| Arousal | 207 | 21.4 | No | 182 | 18.8 |
| Adherence | 595 | 61.5 | I don’t know (missing) | 288 | 29.7 |
| Eudaimonic Project | 155 | 16.0 |  |  |  |
| **Reduced restaurant meals** |  |  | **Stockpiling** |  |  |
| Yes | 323 | 33.3 | Yes | 52 | 5.3 |
| No | 645 | 66.7 | No | 916 | 94.7 |
| **Reduced ethnic restaurant meals** |  |  | **Fresh food** |  |  |
| Yes | 332 | 34.2 | Diminished | 15 | 1.4 |
| No | 636 | 65.8 | Unchanged | 872 | 90.1 |
|  |  |  | Increased | 76 | 7.9 |
|  |  |  | Not buying (missing) | 5 | .6 |
| **Frozen food** |  |  | **Personal care** |  |  |
| Diminished | 13 | 1.2 | Diminished | 10 | 1.0 |
| Unchanged | 867 | 89.6 | Unchanged | 848 | 87.6 |
| Increased | 69 | 7.2 | Increased | 91 | 9.4 |
| Not buying (missing) | 19 | 1.9 | Not buying (missing) | 19 | 2.0 |
| **Canned food** |  |  | **Personal disinfection** |  |  |
| Diminished | 17 | 1.7 | Diminished | 9 | .8 |
| Unchanged | 821 | 84.9 | Unchanged | 735 | 76.0 |
| Increased | 98 | 10.1 | Increased | 185 | 19.2 |
| Not buying (missing) | 32 | 3.3 | Not buying (missing) | 39 | 4.1 |
|  |  |  | **House disinfection** |  |  |
|  |  |  | Diminished | 12 | 1.3 |
|  |  |  | Unchanged | 780 | 80.6 |
|  |  |  | Increased | 142 | 14.7 |
|  |  |  | Not buying (missing) | 34 | 3.4 |

**Table 4.**
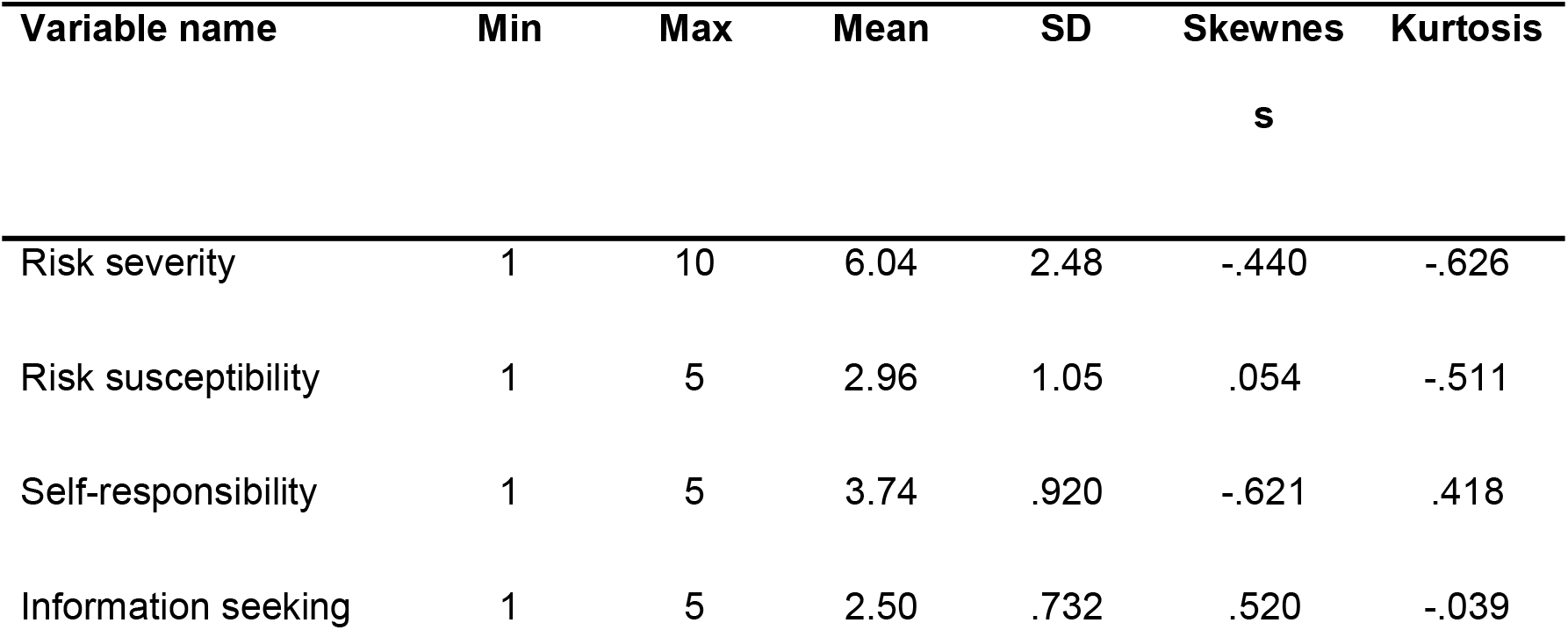
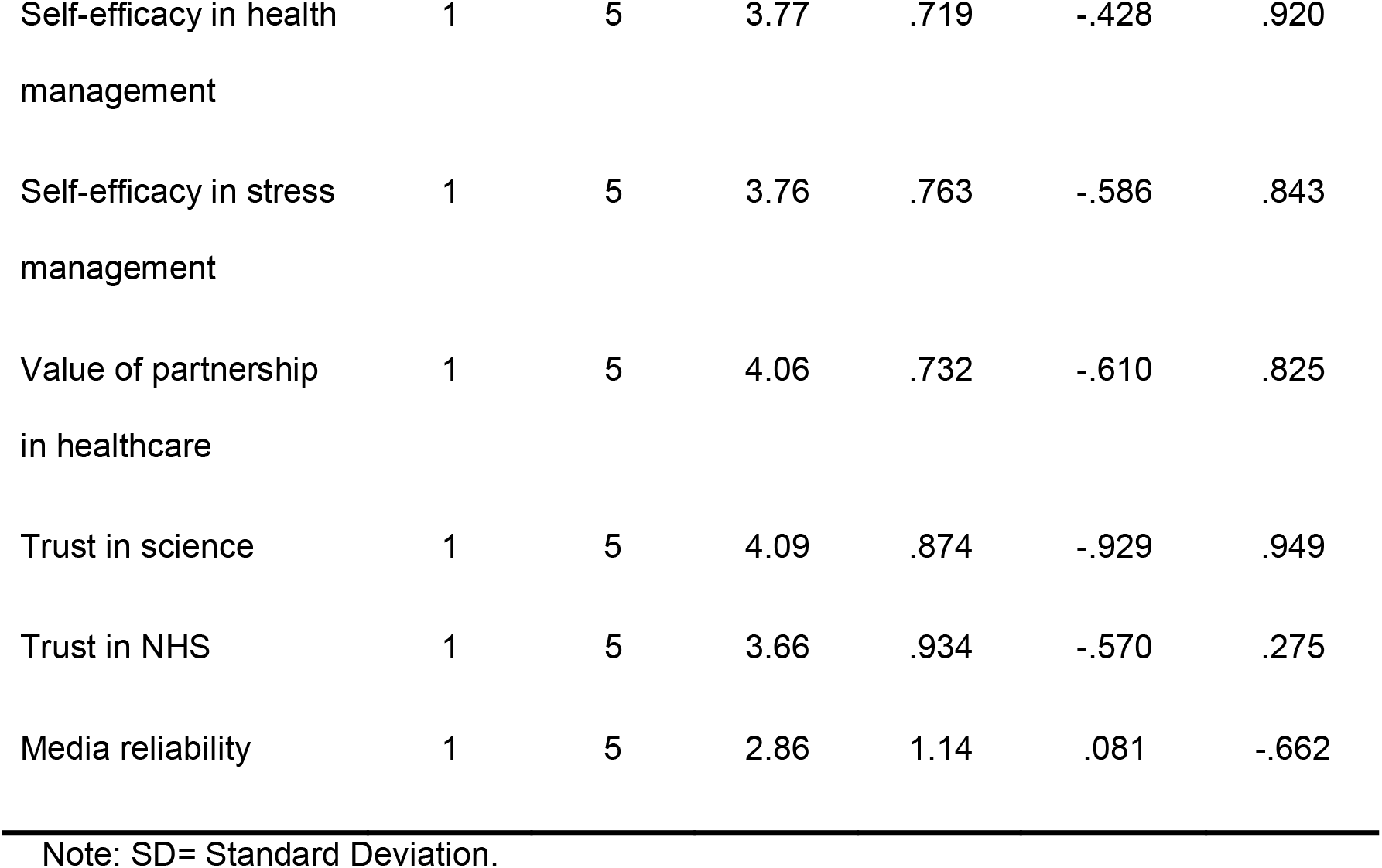
Descriptive statistics for items

Since very few participants resulted being in “Blackout” position, they were grouped together with participants in “Arousal” to facilitate statistical analyses.

### 3.4 Attitudinal response towards to the COVID-19 health emergency

#### 3.4.1 Risk severity

A factorial ANOVA with Risk Severity for the emergency as dependent variable and Health Engagement and “Coming from red zones” as independent variables was carried followed by Tuckey HSD post-hoc tests. TukeyHSD post-hoc test was preferred since it is conservative when there are unequal sample sizes.

Results show a significative main effect of Health Engagement on Risk Susceptibility [F_(2, 1048)_=185.709; p<.001; η^2^_p_=.262]. No other significant main effect or interaction was found. Tukey post-hoc comparisons were carried out to confront the averages of the different Health Engagement levels. In particular, the Arousal group (M=8.00; SD=1.71) was found to be more concerned than both the Adhesion group (M=5.98; SD=2.09) and the Eudaimonic Project group (M=3.51; SD=2.39) with a significance level of 99.9%. Also, the means difference of Adhesion and Eudaimonic Project groups was found to be statistically significant with p<.001.

#### 3.4.2 Risk susceptibility

A factorial ANOVA with Risk Susceptibility as dependent variable and Health Engagement and “Coming from red zones” as independent variables was carried followed by Tuckey HSD post-hoc tests. Results show a significative main effect of Health Engagement on Risk Susceptibility [F_(2, 1040)_=150.890; p<.001; η^2^_p_=.225]. No other significant main effect or interaction was found. Tukey post-hoc comparisons were carried out to confront the averages of the different Health Engagement levels. In particular, the Arousal group (M=3.73; SD=.87) was found to perceive themselves as more at risk than both the Adhesion group (M=2.94; SD=.92) and the Eudaimonic Project group (M=1.97; SD=.906) with a significance of 99.9%. Also the means difference of Adhesion and Eudaimonic Project groups was found to be statistically significant with p<.001.

#### 3.4.3 Orientation towards health management and trust in authorities

A series of univariate Welch’s ANOVAs with Health Engagement as independent variable was carried out followed by Games-Howell post-hoc comparisons to investigate the self-efficacy in health management and different levels of trust in authorities across the Health Engagement levels. Welch’s ANOVA and G-H post-hoc comparisons were preferred over a classic ANOVA approach to provide a more robust method for data analysis (40) since some dependent variables were violating the assumption of homoschedasticity.

Results show a significative main effect of Health Engagement on Self-Responsibility [F_(2, 322.257)_=3.700; p=.026; η^2^=.009], Self-Efficacy in Health Management [F_(2, 339.819)_=57.382; p<.001; η^2^ =.113], Self-Efficacy in Stress Management [F_(2, 355.911)_=16.497; p<.001; η^2^ =.032], Value of Partnership in Healthcare [F_(2, 344.585)_=9.568; p<.001; η^2^=.022], Trust in Science [F_(2, 335.022)_=8.158; p=.001; η^2^=.018], Trust in NHS [F_(2, 337.641)_=9.575; p<.001; η^2^=.021] and Media Reliability [F_(2, 344.288)_=28.664; p<.001; η^2^=.060]. Results of Games-Howell comparisons are reported in Table 5.

**Table 5.** Results of Games-Howell comparisons

| Dependent variables | Engagement Level Comparison |  |  |
| --- | --- | --- | --- |
|  | Arousal-Adhesion | Arousal-Eudaimonic | Adhesion-eudaimonic |
| Self-responsibility | -.162 (.073) | -.274 (.110)* | -.112 (.095) |
| Self-efficacy in health management | -.326 (.057)*** | -.791 (.074)*** | -.465 (.060)*** |
| Self-efficacy in stress management | -.122 (.059) | -.434 (.077)*** | -.312 (.066)*** |
| Value of partnership in healthcare | -.205 (.062)** | -.335 (.078)*** | -.130 (.062) |
| Trust in science | -.218 (.071)** | -.378 (.099)** | -.160 (.081) |
| Trust in NHS | -.245 (.072)** | -.425 (.104)** | -.181 (.091) |
| Media | -.352 (.084)*** | -.911 (.120)*** | -.559 (.107)*** |

Note: values in cells are differences in means. Standard errors are reported in brackets.
Significance is marked with asterisks (\* sig. at $p < .05$ ; \*\* sig. at $p < .01$ ; \*\*\*sig at $p < .001$ ).

### 3.5 Behavioral responses

#### 3.5.1 Information seeking

A Welch’s ANOVA with Health Engagement as independent variable and Information Seeking as dependent was carried out followed by Games-Howell post-hoc comparisons to investigate whether people in different Health Engagement positions show different amounts of media access. Results show a significant main effect of Health Engagement on Information Seeking [F_(2, 334.095)_=29.344; p<.001; η^2^=.064]. G-H comparisons showed that the amount of Information Seeking different significantly among all the different levels: in particular, results showed that people in Arousal search significantly more information (M=2.79; SD=.74) than both people in Adhesion (M=2.47; SD=.68) and Eudaimonic project (M=2.20; SD=.77). The comparison amongst Adhesion and Eudaimonic project resulted significantly different as well.

#### 3.5.2 Consumer habits and purchasing behaviors

Dichotomous variables were used as dependent variables in a series of logistic regressions, in order to understand whether the Health Engagement position is predictive of a change in consumer habits. Risk Susceptibility and Risk Severity were also used as predictors. Wald forward method was selected to automatically exclude non-significant predictors. Health Engagement was used as a categorical variable and hence dummy coded: Eudaimonic Project was used as the 0, the baseline of comparison, for the other two levels. Dependent variables were coded so that “No” was used as the comparison level for “Yes”. Hence, an Odds Ration > 1 should be interpreted as “more likely to answer yes” and vice-versa. Results are reported in Table 6. To assess the association between change in consumer purchase behaviors and different Health Engagement levels a series of contingency tables was created. Pearson’s Chi-square and Fisher’s exact tests were also carried out to reject the null hypothesis that data are randomly distributed across Health Engagement levels. As post-hoc, standardized residuals were inspected: standardized residuals are calculated as the difference between observed and expected counts of a cell divided by an estimate of its standard deviation. Since they are asymptotically normally distributed with a mean of 0 and standard deviation of 1 under the null hypothesis of independence, as a general rule of thumb cells with an absolute value of standard residuals above 2 can be considered to significantly contribute to the general chi-square value (41). For Stockpiling behavior groups were way too unbalanced to proceed with a logistic regression (Yes=5.6%), hence an approach based on contingency tables was preferred. Results are reported in Table 7.

**Table 6.**
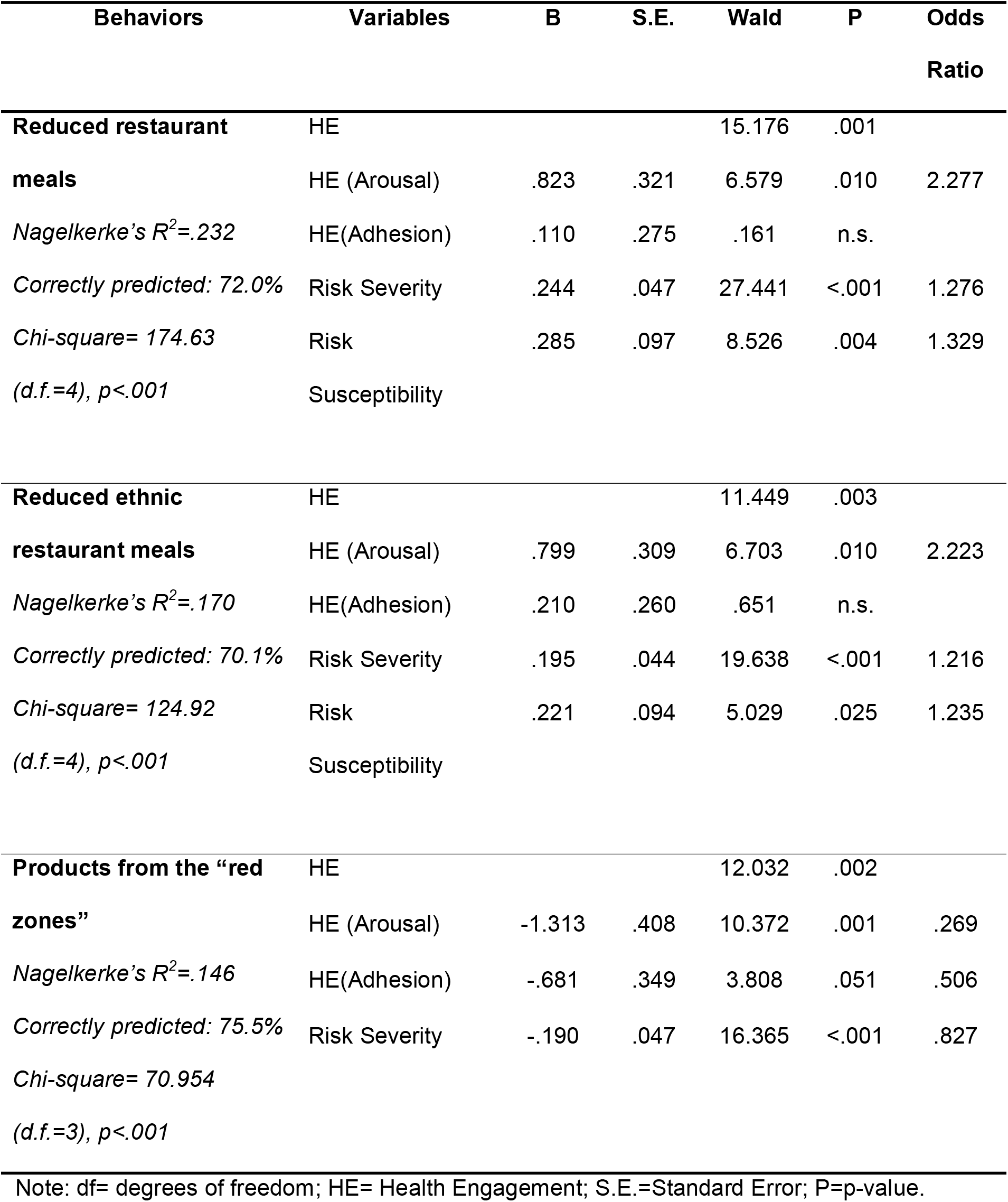
Results of logistic regressions

| Behaviors | Variables | B | S.E. | Wald | P | Odds Ratio |
| --- | --- | --- | --- | --- | --- | --- |
| **Reduced restaurant meals** | HE |  |  | 15.176 | .001 |  |
|  | HE (Arousal) | .823 | .321 | 6.579 | .010 | 2.277 |
| *Nagelkerke's $R^2$=.232* | HE(Adhesion) | .110 | .275 | .161 | n.s. |  |
| *Correctly predicted: 72.0%* | Risk Severity | .244 | .047 | 27.441 | <.001 | 1.276 |
| *Chi-square= 174.63* | Risk | .285 | .097 | 8.526 | .004 | 1.329 |
| *(d.f.=4), $p$<.001* | Susceptibility |  |  |  |  |  |
| **Reduced ethnic restaurant meals** | HE |  |  | 11.449 | .003 |  |
|  | HE (Arousal) | .799 | .309 | 6.703 | .010 | 2.223 |
| *Nagelkerke's $R^2$=.170* | HE(Adhesion) | .210 | .260 | .651 | n.s. |  |
| *Correctly predicted: 70.1%* | Risk Severity | .195 | .044 | 19.638 | <.001 | 1.216 |
| *Chi-square= 124.92* | Risk | .221 | .094 | 5.029 | .025 | 1.235 |
| *(d.f.=4), $p$<.001* | Susceptibility |  |  |  |  |  |
| **Products from the “red zones”** | HE |  |  | 12.032 | .002 |  |
|  | HE (Arousal) | -1.313 | .408 | 10.372 | .001 | .269 |
| *Nagelkerke's $R^2$=.146* | HE(Adhesion) | -.681 | .349 | 3.808 | .051 | .506 |
| *Correctly predicted: 75.5%* | Risk Severity | -.190 | .047 | 16.365 | <.001 | .827 |
| *Chi-square= 70.954* |  |  |  |  |  |  |
| *(d.f.=3), $p$<.001* |  |  |  |  |  |  |

Note: df= degrees of freedom; HE= Health Engagement; S.E.=Standard Error; P=p-value.

**Table 7.**
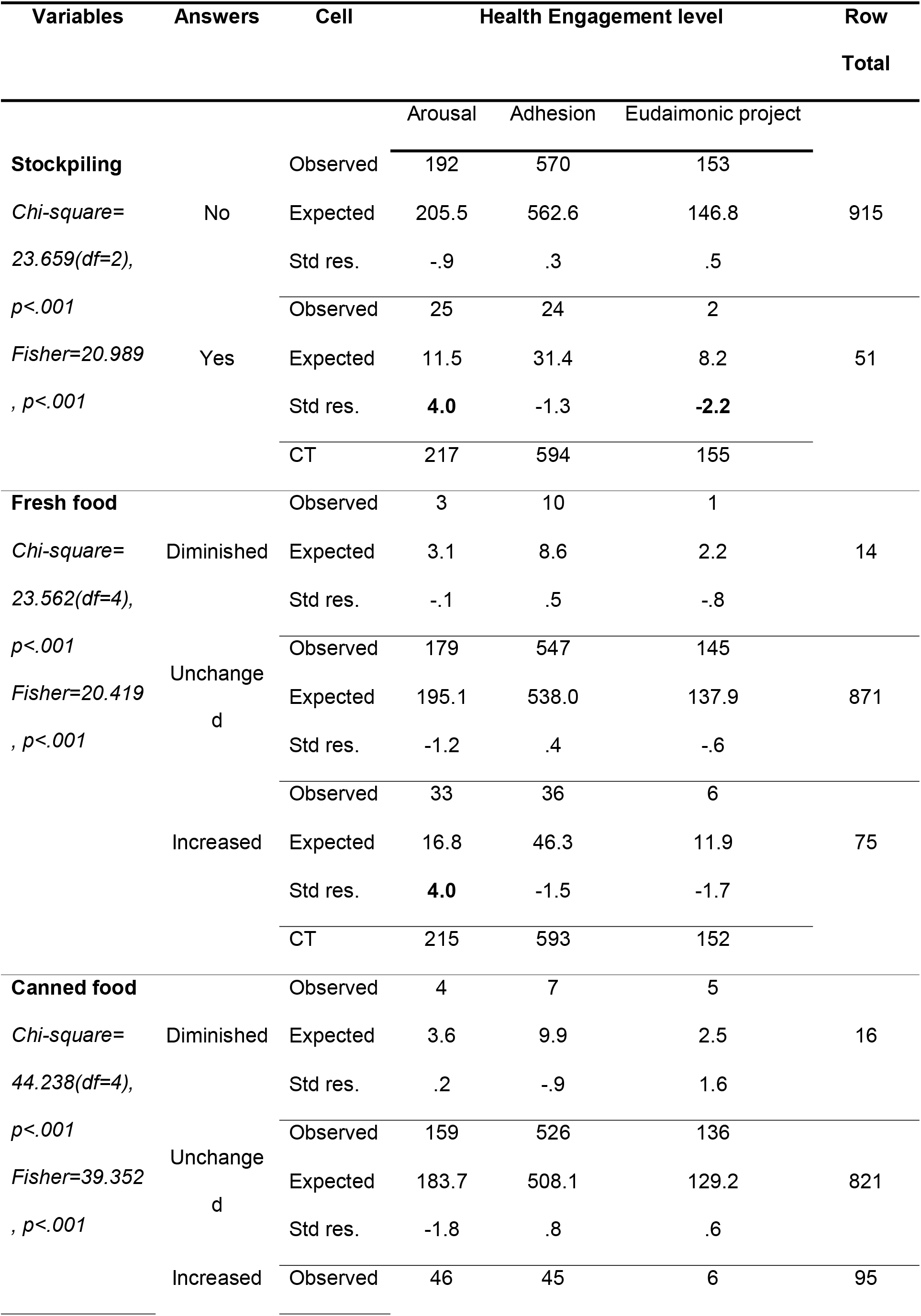
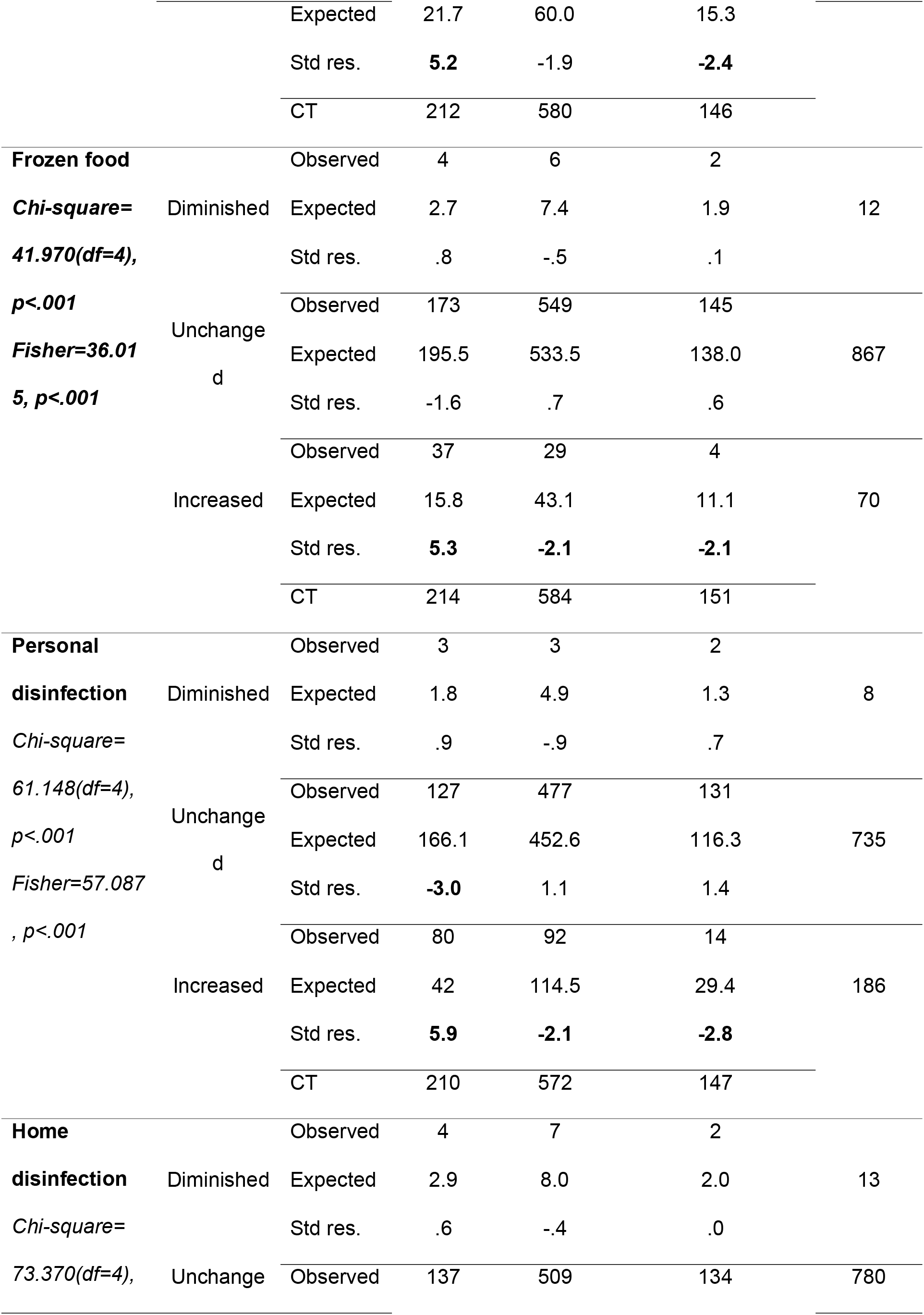
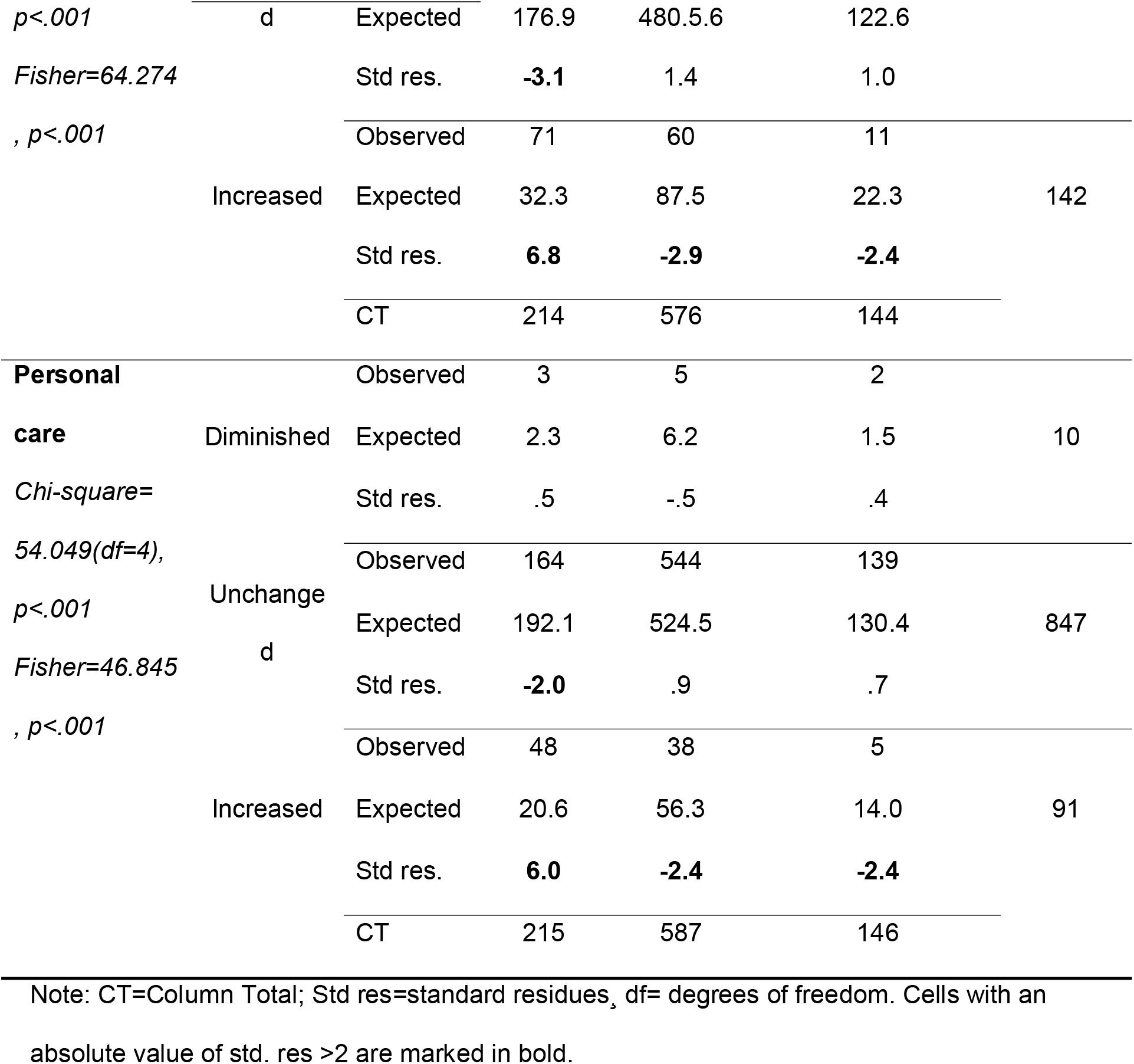
Results of contingency tables

## 4. Discussion

At the end of February 2020 the diffusion of the COVID-19 epidemics in northern Italy has forced health authorities to embrace restrictive preventive measures that impacted on Italian citizens’ daily habits and consumption behaviors. Enforcing compliance to such measures was crucial at that time in order to enhance their effectiveness and to sustain the sustainability of the healthcare system. However, this sudden change determined huge reactions by Italian citizens: many of them experienced panic and enacted maladaptive behaviors. In this scenario, the Italian citizens’ reactions to the COVID-19 emergency measures, from the scientific perspective, is an interesting and unique platform to demonstrate the value of making citizens engaged and actual partner of the healthcare system to safeguard both individuals’ and collective health. Therefore, we consider the current COVID-19 outbreak in Italy as a valuable “testing ground” for consumer education initiatives aimed at sustaining their health engagement and compliance to the prescribed behavioral changes.

Existing research has focused on demographic and immutable factors and subjective that influence how people are likely to behave in a pandemic (42–44). Furthermore, previous research on responses to pandemics has been largely a-theoretical (8). Therefore, all these studies provide valuable insights into how different segment of the population are likely to respond, but does not tell us why they respond in this way.

Therefore, the current study adopted the theoretical lenses of the Patient Health Engagement Model to explain – from a psychosocial perspective - people responses through the first wave of the COVID-19 pandemic in Italy. This theory states that individuals are more or less akin to change their behaviors according to their own subjective perceptions about the role (more or less active) the might play in their health and care (32).

The Patient Health Engagement Model (PHE) provides a potentially useful framework for understanding how people will respond to health threats such as pandemics and related prescribed preventive measures imposed by healthcare authorities. The PHE model proposes that people’s adaptive behavioral and emotional responses to protect themselves from a health threat is influenced by the level of health engagement – that is a progressive reframing of individuals’ own role within the healthcare system (i.e. from passive user of services to active partner of the healthcare system) (32). In this study we employed and validated a revised version of the PHE-s^®^ to measure citizens’ health engagement. This revised version showed good psychometric properties for our representative sample.

According to the study results, Italian citizens seems to be more concerned for the health emergency than not, even though not extremely worried (on a scale from 1 to 10, the average is around 6) and not feeling exceedingly at risk of being infected (the 5-point Likert shows a normaloid distribution with mean around the central point), confirming previous studies in other similar settings (6,45,46). Nevertheless, it is important to notice how different Health Engagement profiles, are associated with different levels of both perceived risk severity and susceptibility: indeed, less engaged people (rated as in “Blackout” and “Arousal”) show significantly higher levels of perceived susceptibility to and perceived risk of the infection when compared with high engaged ones (rated as in “Adhesion” and “Eudaimonic Project”), regardless of the geographical area of origin (“red zone” or not), which surprisingly didn’t result to be associated with different levels of susceptibility and severity. This seems to testify that people differ in their ability to psychologically master their worries related to the COVID-19 epidemic, and this explains consequent more or less adherence to the change in behaviors imposed by the health authorities. This interpretation is confirmed also by the fact that people with different levels of Health Engagement show different attitudinal responses to the emergency: in particular, when compared with people with higher levels of health engagement, less engaged people are less trusting scientific and healthcare authorities, they feel less self-effective in managing their own health - both in normal conditions and under stress - and less prone to cooperate with healthcare professionals. These results are confirmative of previous studies on Influenza A(H1N1) which demonstrated that if perceived severity and susceptibility are high but response and self-efficacy are low, maladaptive responses (e.g. denying the existence of a threat) are likely to ensue (47,48). The perceived self-efficacy in health management and a sentiment of mistrust towards authorities may actually help in understanding why a less engaged person feels more concerned and worried for the new COVID-19 emergency: they seek more information, potentially exposing themselves to fake or over-hyped news, since they are also prone to feel that news regarding the emergency are reliable; nevertheless, they mistrust scientific research and the capacity of the NHS to cope with the pandemic and feel less capable of taking care of themselves. Furthermore, low levels of Health Engagement may testify that people do not consider themselves ready to be active partners of the healthcare systems, being more focused on their own health interests and need and not incline to collaborate and trust the healthcare system to achieve a common public health goal.

Health Engagement also seems to be a predictor of behavioral responses to the emergency. Generally speaking, a substantial part of our sample reported a change in their habits: one out of three Italian citizens reported to have reduced meals outside and/or meals in ethnic restaurants, while 20% declared that they would not buy products coming from “red zones”. Indeed, while Risk Severity and Risk Susceptibility are clearly strong predictors, logistic models show that people with lower levels of Health Engagement have more than twice the chances than people with higher level of health engagement to either have reduced meals or ethnic meals outside home. It’s important to notice that data have been collected in a moment where the emergency was still away from its peak and guidelines were not forbidding people from moving freely and have meals in restaurants. These results could be interpreted in line with previous studies that underlined as when unknown diseases are thought to be lethal, people are incline to blame the outbreaks on someone, or some group of people, who live outside of their own social sphere, as a mechanism to cope with fear and risk perception (49). In this research, it appears clear as this form of “moral panic” (50) had an halo effect also on products and restaurants people naively thought to be guilty in Covid-19’s spread, or that were related to the “infected zone”. Such lay interpretation of disease transmission, together with the difficulty of finding reliable information in a first phase of health emergency, has an impact on people’s habits and consumptions, and clear consequences on local enterprises’ economy. A similar case occurred with the H5N1 Avian Influenza on food consumption, when the poultry industry has suffered severe losses due to a sort of “halo effect” in consumer perception of risk, even after the emergency (51,52).

Despite these results, for what concerns buying behaviors, our data show that generally most people didn’t actually change their habits, in line with other studies (53): most people didn’t stockpile goods nor increased the purchase of the goods we have considered in our survey. Nevertheless, crosstabs show that amongst those who stockpiled goods and increased the purchase of food (fresh, frozen or canned) and disinfection products (in particular regarding home disinfection) there is a significantly higher presence of lower engaged consumers. This evidence in line with other studies (54–56) that showed how personal reaction of the critical event can feed behavioral changes, with many people enacting significant changes in their consumption behaviors like anticipating the purchasing of goods (57,58). As food consumption is recognized as a primary need for the individuals, it is strongly influenced by the subjective interpretation of risk and the possible scarcity (59). For this reason, these results appear interesting in giving a sense to how people orient their food purchase in the case of emergency in relation to their engagement level. Furthermore, it appears evident how people with low level of Health Engagement, not being psychologically ready to consider the social and public health consequences of their conduct, appear more focused on their own health interests and less keen to rely on health authorities’ guidelines to orient their behaviors. For instance, the behavior of stockpile goods enacted by the less engaged Italian citizens had a negative organizational impact on food supplies which further compromised the delicate situation of the Italian population. Furthermore, the overcrowding at superstores in the situation of COVID-19 epidemics was highly counterproductive and contributed to spread the risk of contagion.

## 5. Limitations and future studies

The study measured a specific population’s views at a specific point in time; their beliefs and attitudes reflect the information available at the time and therefore are not stable. Second, results were self-reported. Measurement errors and social desirability bias may exist although the study was anonymous.

Future research should test the Patient Health Engagement Model as a predictor of particular behaviors such as hand-hygiene or facial masks usage. In addition, it is important to carry out further behavioral research where actual behavior can be measured and not only self-reported.

## 6. Practical implications

This study has provided evidence about the role of health engagement as a determinant of citizens’ behavioral change which is key for controlling the spread of pandemic disease, and described a conceptual framework – i.e. the Patient Health Engagement Model - in which to better understand these behaviors. In sum, the study shows that health engagement levels are predictive of different responses, both affective and behavioral: playing an active role in health management is associated with a higher chance of enacting specific behaviors. In particular, the psychological readiness to assume a proactive role in one own health prevention is explicative of the individuals’ tendency to be more or less able to comply to health authorities’ prescription and to perceive themselves has main responsible for their own health and the health of their community. Furthermore, the psychological readiness to engage in health results in a crucial factor for explaining the different way in which individuals can cope with their worries for a health emergency. The findings suggest that intervention studies should focus on particular groups and on raising their levels of engagement to increase the effectiveness of educational initiatives devoted to promoting preventive behaviors. Communication strategies should maximize their impact by targeting messages according to the health engagement levels of citizens. For instance, in order to improve the levels of engagement of citizens in a “psychological blackout”, reassuring messages, aimed at sustaining the emotional elaboration of the emergency and related worries would be particularly needed. Citizens in a situation of “psychological adherence” would need positive stories of other persons who succeeded in adhere to the prescribed containment measures in order to enhance their motivation to stay engaged. Finally, people in the position of “eudaimonic projects”, who were able to develop a new sense of normality despite the sanitary emergency, can be involved in peer-to-peer communication initiatives becoming advocates for the correct engagement in adhere to the prescribed measures to face the COVID-19 epidemic. Furthermore, fostering the psychological readiness to get engaged in health prevention appears to be a crucial goal for educational and communication initiatives in the situation of a health emergency. Carrying out this work now will be invaluable in preparing for this and future pandemics.

## Data Availability

Data are available upon request to the authors

## Declarations

### Ethics approval and consent to participate

Ethical approval was obtained from the institutional review board at Catholic University of Milan (IRB#2019-12).

### Consent for publication

Not applicable

### Availability of data and materials

The authors are not authorized to share the dataset publicly due to potentially sensitive data. However it is available from the corresponding author on reasonable request.

### Authors’ contributions

GG conceptualized and supervised the writing of the paper. SB and MS drafted the paper. LP, GC and AB conducted the statistical analysis presented in the paper and provided assistance in writing methods and results. GG, SB, MS, LP, GC, AB and EL all equally contributed to the design of the survey, data interpretation and discussion of intellectual content. All authors read and approved the final manuscript.

## References

1. Godoy P, Castilla J, Delgado-Rodríguez M, Martín V, Soldevila N, Alonso J, et al. Effectiveness of hand hygiene and provision of information in preventing influenza cases requiring hospitalization. Prev Med (Baltim). 2012;54(6):434–9.

2. Ferguson NM, Cummings DAT, Fraser C, Cajka JC, Cooley PC, Burke DS, et al. Strategies for mitigating an influenza pandemic. Nature. 2006;

3. Fenichela EP, Castillo-Chavezb C, Ceddiac MG, Chowellb G, Gonzalez Parrae PA, Hickling GJ, et al. Adaptive human behavior in epidemiological models. Proc Natl Acad Sci U S A. 2011;

4. Funk S, Salathé M, Jansen VAA. Modelling the influence of human behaviour on the spread of infectious diseases: A review. Journal of the Royal Society Interface. 2010.

5. Fenichel EP, Kuminoff N V., Chowell G. Skip the Trip: Air Travelers’ Behavioral Responses to Pandemic Influenza. PLoS One. 2013;8(3).

6. Leppin A, Aro AR. Risk perceptions related to SARS and avian influenza: Theoretical foundations of current empirical research. Int J Behav Med. 2009;16(1):7–29.

7. Kass NE, Otto J, O’Brien D, Minson M. Ethics and severe pandemic influenza: Maintaining essential functions through a fair and considered response. Biosecurity and Bioterrorism. 2008;

8. Bish A, Michie S. Demographic and attitudinal determinants of protective behaviours during a pandemic: A review. Br J Health Psychol. 2010;15(4):797–824.

9. Jin Y, Pang A, Cameron GT. Toward a Publics-Driven, Emotion-Based Conceptualization in Crisis Communication: Unearthing Dominant Emotions in Multi-Staged Testing of the Integrated Crisis Mapping (ICM) Model. J Public Relations Res. 2012;

10. Zhang Y, Yang H, Cheng P, Luqman A. Predicting consumers’ intention to consume poultry during an H7N9 emergency: an extension of the theory of planned behavior model. Hum Ecol Risk Assess. 2020;

11. Mossong J, Hens N, Jit M, Beutels P, Auranen K, Mikolajczyk R, et al. Social contacts and mixing patterns relevant to the spread of infectious diseases. PLoS Med. 2008;

12. Lau JTF, Griffiths S, Au DWH, Choi KC. Changes in knowledge, perceptions, preventive behaviours and psychological responses in the pre-community outbreak phase of the H1N1 epidemic. Epidemiol Infect. 2011;139(1):80–90.

13. Brug J, Aro AR, Richardus JH. Risk perceptions and behaviour: Towards pandemic control of emerging infectious diseases: Iional research on risk perception in the control of emerging infectious diseases. Int J Behav Med. 2009;16(1):3–6.

14. Poletti P, Ajelli M, Merler S. The effect of risk perception on the 2009 H1N1 pandemic influenza dynamics. PLoS One. 2011;

15. Smith RD. Responding to global infectious disease outbreaks: Lessons from SARS on the role of risk perception, communication and management. Soc Sci Med. 2006;63(12):3113–23.

16. De Zwart O, Veldhuijzen IK, Elam G, Aro AR, Abraham T, Bishop GD, et al. Perceived threat, risk perception, and efficacy beliefs related to SARS and other (emerging) infectious diseases: Results of an international survey. Int J Behav Med. 2009;

17. Brug J, Aro AR, Oenema A, De Zwart O, Richardus JH, Bishop GD. SARS risk perception, knowledge, precautions, and information sources, the Netherlands. Emerg Infect Dis. 2004;

18. Vartti AM, Oenema A, Schreck M, Uutela A, De Zwart O, Brug J, et al. SARS knowledge, perceptions, and behaviors: A comparison between finns and the dutch during the SARS outbreak in 2003. Int J Behav Med. 2009;

19. Cowling BJ, Ng DMW, Ip DKM, Liao Q, Lam WWT, Wu JT, et al. Community Psychological and Behavioral Responses through the First Wave of the 2009 Influenza A(H1N1) Pandemic in Hong Kong. J Infect Dis. 2010;202(6):867–76.

20. Jones JH, Salathé M. Early assessment of anxiety and behavioral response to novel swine-origin influenza a(H1N1). PLoS One. 2009;4(12):2–9.

21. Rubin GJ, Amlôt R, Page L, Wessely S. Public perceptions, anxiety, and behaviour change in relation to the swine flu outbreak: Cross sectional telephone survey. BMJ. 2009;339(7713):156.

22. Chapman GB, Coups EJ. Predictors of influenza vaccine acceptance among healthy adults. Prev Med (Baltim). 1999;

23. Ritvo P, Wilson K, Willms D, Upshur R. Vaccines in the public eye. Nature Medicine. 2005.

24. Ibuka Y, Chapman GB, Meyers LA, Li M, Galvani AP. The dynamics of risk perceptions and precautionary behavior in response to 2009 (H1N1) pandemic influenza. BMC Infect Dis. 2010;

25. Coulter A. Patient engagement-what works? J Ambul Care Manage. 2012;

26. Graffigna G, Barello S, Bonanomi A, Lozza E. Measuring patient engagement: Development and psychometric properties of the patient health engagement (PHE) scale. Front Psychol. 2015;6(MAR):1–10.

27. Barello S, Graffigna G, Vegni E. Patient Engagement as an Emerging Challenge for Healthcare Services: Mapping the Literature. Nurs Res Pract. 2012;

28. Gruman J, Rovner MH, French ME, Jeffress D, Sofaer S, Shaller D, et al. From patient education to patient engagement: Implications for the field of patient education. Patient Educ Couns. 2010;

29. Graffigna G, Barello S, Bonanomi A. The role of Patient Health Engagement model (PHE-model) in affecting patient activation and medication adherence: A structural equation model. PLoS One. 2017;12(6).

30. Graffigna G, Barello S, Bonanomi A, Riva G. Factors affecting patients’ online health information-seeking behaviours: The role of the Patient Health Engagement (PHE) Model. Patient Educ Couns. 2017;

31. Hill AM, Etherton-Beer C, Haines TP. Tailored Education for Older Patients to Facilitate Engagement in Falls Prevention Strategies after Hospital Discharge-A Pilot Randomized Controlled Trial. PLoS One. 2013;

32. Graffigna G, Barello S. Spotlight on the patient health engagement model (PHE model): A psychosocial theory to understand people’s meaningful engagement in their own health care. Patient Preference and Adherence. 2018.

33. Griffin RJ, Dunwoody S, Neuwirth K. Proposed model of the relationship of risk information seeking and processing to the development of preventive behaviors. In: Environmental Research. 1999.

34. Wright BD, Linacre JM, Gustafson JE, Martin-Lof P. Reasonable mean-square fit values. Rasch Meas Trans. 1994;8(3):370.

35. Bonanomi A, Cantaluppi G, Nai Ruscone M, Osmetti SA. A new estimator of Zumbo’s Ordinal Alpha: a copula approach. Qual Quant [Internet]. 11 maggio 2015;49(3):941–53. Available at: http://link.springer.com/10.1007/s11135-014-0114-8

36. Gliem JA, Gliem RR. Calculating, Interpreting, and Reporting Cronbach’s Alpha Reliability Coefficient for Likert-Type Scales. In: Midwest Research-to-Practice Conference in Adult, Continuing, and Community Education. Columbus, OH, USA: The Ohio State University press; 2003.

37. Bentler PM. Comparative fit indexes in structural models. Psychol Bull. 1990;107(2):238.

38. Jöreskog KG, Sörbom D, Lisrel VI. Analysis of linear structural relationship by maximum likelihood. Chicago: Scientific Software; 1984.

39. MacCallum RC, Browne MW, Sugawara HM. Power analysis and determination of sample size for covariance structure modeling. Psychol Methods. 1996;1(2):130.

40. Delacre M, Leys C, Mora YL, Lakens D. Taking Parametric Assumptions Seriously: Arguments for the Use of Welch’s F-test instead of the Classical F-test in One-Way ANOVA. Int Rev Soc Psychol. 2019;

41. Haberman SJ. The Analysis of Residuals in Cross-Classified Tables. Biometrics. 1973;

42. J. F, P. S, K. MC. Gender, race, and perception of environmental health risks. Risk Anal. 1994;

43. Balinska M, Rizzo C. Behavioural responses to influenza pandemics. PLoS Currents. 2009.

44. Tooher R, Collins JE, Street JM, Braunack-Mayer A, Marshall H. Community knowledge, behaviours and attitudes about the 2009 H1N1 Influenza pandemic: A systematic review. Influenza Other Respi Viruses. 2013;

45. Kristiansen IS, Halvorsen PA, Gyrd-Hansen D. Influenza pandemic: Perception of risk and individual precautions in a general population. Cross sectional study. BMC Public Health. 2007;

46. Balkhy HH, Abolfotouh MA, Al-Hathlool RH, Al-Jumah MA. Awareness, attitudes, and practices related to the swine influenza pandemic among the Saudi public. BMC Infect Dis. 2010;

47. Kok G, Jonkers R, Gelissen R, Meertens R, De Zwart O, Schealma H. Behavioural intentions in response to an influenza pandemic. BMC Public Health. 2010;10.

48. Ruiter RAC, Abraham C, Kok G. Scary warnings and rational precautions: A review of the psychology of fear appeals. Psychol Heal. 2001;16(6):613–30.

49. McCauley M, Minsky S, Viswanath K. The H1N1 pandemic: Media frames, stigmatization and coping. BMC Public Health. 2013;13(1):1–16.

50. Muzzatti SL. Bits of falling sky and global pandemics: Moral panic and Severe Acute Respiratory Syndrome (SARS). Illness Crisis and Loss. 2005.

51. Akben E, Özertan G, Spaulding AD, Saghaian SH. Consumer responses of the H5N1 Avian Influenza: The case of Turkey. Econ Bull. 2008;4(15).

52. de Krom MPMM, Mol APJ. Food risks and consumer trust. Avian influenza and the knowing and non-knowing on UK shopping floors. Appetite. 2010;55(3):671–8.

53. Van D, McLaws ML, Crimmins J, MacIntyre CR, Seale H. University life and pandemic influenza: Attitudes and intended behaviour of staff and students towards pandemic (H1N1) 2009. BMC Public Health. 2010;10.

54. Goodwin R, Haque S, Neto F, Myers LB. Initial psychological responses to Influenza A, H1N1 ("Swine flu"). BMC Infect Dis. 2009;9:166.

55. Bults M, Beaujean DJMA, De Zwart O, Kok G, Van Empelen P, Van Steenbergen JE, et al. Perceived risk, anxiety, and behavioural responses of the general public during the early phase of the Influenza A (H1N1) pandemic in the Netherlands: Results of three consecutive online surveys. BMC Public Health. 2011;

56. Kamate SK, Agrawal A, Chaudhary H, Singh K, Mishra P, Asawa K. Public knowledge, attitude and behavioural changes in an Indian population during the Influenza A (H1N1) outbreak. J Infect Dev Ctries. 2010;

57. Fischhoff B, De Bruin WB, Perrin W, Downs J. Travel risks in a time of terror: Judgments and choices. Risk Anal. 2004;

58. Kim HK, Niederdeppe J. The Role of Emotional Response during an H1N1 Influenza Pandemic on a College Campus. J Public Relations Res. 2013;25(1):30–50.

59. Gstraunthaler T, Day R. Avian influenza in the UK: Knowledge, risk perception and risk reduction strategies. Br Food J. 2008;110(3):260–70.

